# Generalizable Prediction of Alzheimer Disease Pathologies with a Scalable Annotation Tool and an High-Accuracy Model

**DOI:** 10.1101/2025.02.03.25321603

**Authors:** Vivek Gopal Ramaswamy, Monika Ahirwar, Gennadi Ryan, Brittany N. Dugger, Osama Al Dalahmah, Maxim Signaevsky, Dushyant P. Purohit, Vahram Haroutunian, Steven Finkbeiner

## Abstract

Characterizing the cardinal neuropathologies in Alzheimer disease (AD) can be laborious, time consuming, and susceptible to intra- and inter-observer variability. The lack of high throughput unbiased approaches to reliably assess neuropathology hampers efforts to use pathology as a means to link clinical features of AD to molecular pathogenesis in the ever-growing datasets of persons with AD. To remove this roadblock, we designed an annotation tool in addition to a computational pipeline to analyze digital microscopic images of postmortem tissue from persons with AD in a fully automated and unbiased manner in only a fraction of the time taken with conventional approaches and allows neuropathological analyses and lesion quantification at multiple scales. The pipeline includes a Mask Regional-Convolutional Neural Network (Mask R-CNN) we trained to detect, classify, and segment different types of amyloid. To establish ground truth for training and validation, we utilized an existing open source platform, QuPath, and developed a tool to collect consensus annotations of neuropathology experts. The Mask R-CNN identified amyloid pathology in samples (with accuracy: 94.6%, F1: 87.7%, Dice: 81.8%) unrelated to the training dataset, indicating that it detects generalizable pathology features. Its quantitative measurements of amyloid pathology on 298 samples correlated with the severity of AD neuropathology assessed by experts and neuropathologists (CERAD ratings) and estimates of cognitive compromise (Clinical Dementia Ratings (CDR)). Our computational pipeline should enable rapid, unbiased, inexpensive, quantitative, and comprehensive neuropathological analysis of large tissue collections and integration with orthogonal clinical and multi-omic measurements.

## Introduction

Alzheimer disease (AD) is the most common adult onset neurodegenerative disease and is characterized by a loss of memory, executive function, and behavioral changes^1,2^. The two diagnostic histopathological hallmarks of AD are the abnormal deposition of amyloid β, a cleavage product of the APP protein, and the presence of neurofibrillary tangles (NFTs) of hyperphosphorylated tau.

Conventionally, amyloid pathology in human brain tissue has been assessed with immunolabeling, and/or histochemical staining, and visual microscopic inspection by expert neuropathologists. In 1991, the Consortium to Establish a Registry for Alzheimer Disease (CERAD) proposed histopathologic diagnostic criteria for AD using a semi-quantitative score of the density of neuritic plaques in the most severely affected regions of the isocortex, and the age at death of the individual, to obtain an age-adjusted plaque score^3^. This semi-quantitative approach to plaque quantification continues, and the specificity of the neuropathological diagnosis of AD has been improved by incorporating Braak and Braak staging of NFTs^4^. New approaches to quantify amyloid β plaques have been developed that utilize machine learning and computer vision. Several studies have demonstrated that deep learning algorithms can achieve human-level accuracy in identifying amyloid β pathology^5,6,7,8^, but most of these models have been tested on relatively small datasets (n < 131). These proof-of-concept studies highlight the potential of machine learning in this domain but may not fully capture the variability and robustness required for broader clinical applications^9^.

Despite the success cited above, many challenges remain. First, digitization of pathology slides creates enormous electronic files, typically ranging from 1GB to 4GB^10^ on average, which are difficult to store and process using machine learning pipelines. Whole slide images (WSI) vary in stain intensity, tissue preparation and slide scanning techniques, complicating analyses. To ingest the imaging data, WSIs are often divided into smaller crops that are easier to compute^6^. But the detection and interpretation of neuropathology depends critically on context. If the crops are too small, some pathology could be overlooked or misdiagnosed^11^. Second, the success of supervised machine learning approaches depends critically on the creation of well-annotated training datasets to establish the ground truth. The quality of ground truth determines and limits the accuracy and generalizability of algorithms produced by the trained machine learning networks^12^. Training datasets are typically generated by experts who hand-annotate examples of characteristic pathology versus background and artifacts in digitized images. But annotations by experts are visual-based and can be subjectively biased^13^. To expedite annotation, many methods ^6,13^ utilize thresholding techniques to detect candidate plaques, which are then sent to annotators for classification. However, this approach often depends on the specific stain, its intensity, and may lack scalability.

There are several tools available to assist in collecting annotations from multiple experts like SuperAnnotate^14^, but to our knowledge, there is no open-source tool that facilitates collecting annotations from different experts and neuropathologists, quantifying consensus, and qualifying ground truth training data. As a result, there is a dearth of studies on generalizability of algorithms to detect AD neuropathology beyond the datasets used to train them, although there have been studies which have validated certain algorithms^15,9^, Fourth, most existing models are either standalone classifiers or segmentation models, typically trained for the detection or segmentation of amyloid pathologies. This separation can limit the models’ ability to perform comprehensive tasks such as simultaneous classification and segmentation, reducing their utility in extracting detailed attributes like plaque size, texture, or diameter, which could enable deeper and more nuanced analysis of these plaques. Additionally, reproducing existing deep learning models is challenging because they are often housed in GitHub repositories, requiring users to have a certain level of expertise to navigate extensive documentation and install various software packages to test the model ^16^.

Here, we present a scalable, open-source platform that addresses most of these problems. Our platform includes an innovative tool for collecting consensus-driven annotations from multiple annotators/experts/neuropathologists, ensuring more accurate and reproducible ground truth data. Our computational pipeline processes 298 whole slide images (WSIs) in a high-throughput manner, requiring only 20 minutes per WSI, consumes 93% less storage and uses a training dataset 15-fold smaller than traditional approaches ^6, 17, 8^ with a single GPU (NVIDIA RTX A4000). Our model simultaneously classifies, segments, and quantifies amyloid β pathologies while extracting additional attributes such as size, texture, and diameter, enabling more robust and scalable analysis. This approach offers a nearly infinite dynamic range for quantitative measures, leveraging the advantages of a ratio scale over the CERAD-type ordinal scale. Furthermore, our model demonstrates strong generalizability across independent datasets and shows a strong correlation with the ordinal amyloid plaque scoring system of CERAD, a key metric for general and regional Alzheimer disease neuropathology severity. The model is deployed as an API (Application Programming Interface) on a GPU cloud and integrated with QuPath through an easy-to-use interface plugin. This setup makes the model accessible to all novices and experts, promoting the broader adoption of machine learning in both research and clinical practice.

## Results

### Expanding Context Window and Using Regional Proposal Networks Reduces Slide Processing Time 5-Fold

WSIs are very large files (100,000 × 100,000 pixels), and batch processing large sets of WSIs can cause premature job termination due to memory overflow errors^18^. WSIs are also too large to be directly ingested into deep learning (DL) networks, so they are typically divided into smaller crops, often 256 x 256 pixels. While this approach makes the data computationally manageable, it can compromise accuracy, as the detection and interpretation of neuropathology depend heavily on contextual information. If the crops are too small, pathologies may be missed or misinterpreted^11,19^.

To optimize throughput and expand the context window, we increased the tile size to 1024 × 1024 pixels from 256 x 256 (Fig. 1, Step 2). To avoid wasting computational time analyzing crops that lack important features, we first preprocessed the WSIs using Otsu thresholding^20^ to get only the foreground pixels corresponding to the tissue area (Supplementary Fig. 3). The coordinates of crops that lie inside the tissue area were saved as a numpy file consuming around 100 Kb of space. This method saved 93% of storage compared to storing all 1024 × 1024 crops, which consumed around 1.5Mb. Next, we added regional proposal network (RPN)^21^ to the architecture of the convolutional neural network (CNN), thus termed Mask R-CNN^22^. The RPN slides a small network window over the convolutional feature map put out by the last layer of the CNN, and determines the probability the window contains an object (Fig.1, Step 4; Supplementary Fig. 4, Step 2). The proposed regions with a high likelihood of containing objects of interest are then fully analyzed by the CNN (Fig. 1, Steps 5-6; Supplementary Fig. 4, Step 2) while low-likelihood cases are ignored. This approach accelerates processing time per image crop from 0.2 seconds to 10 milliseconds^21^, resulting in a 20-fold reduction in computational power required per image.

**Figure 1.**
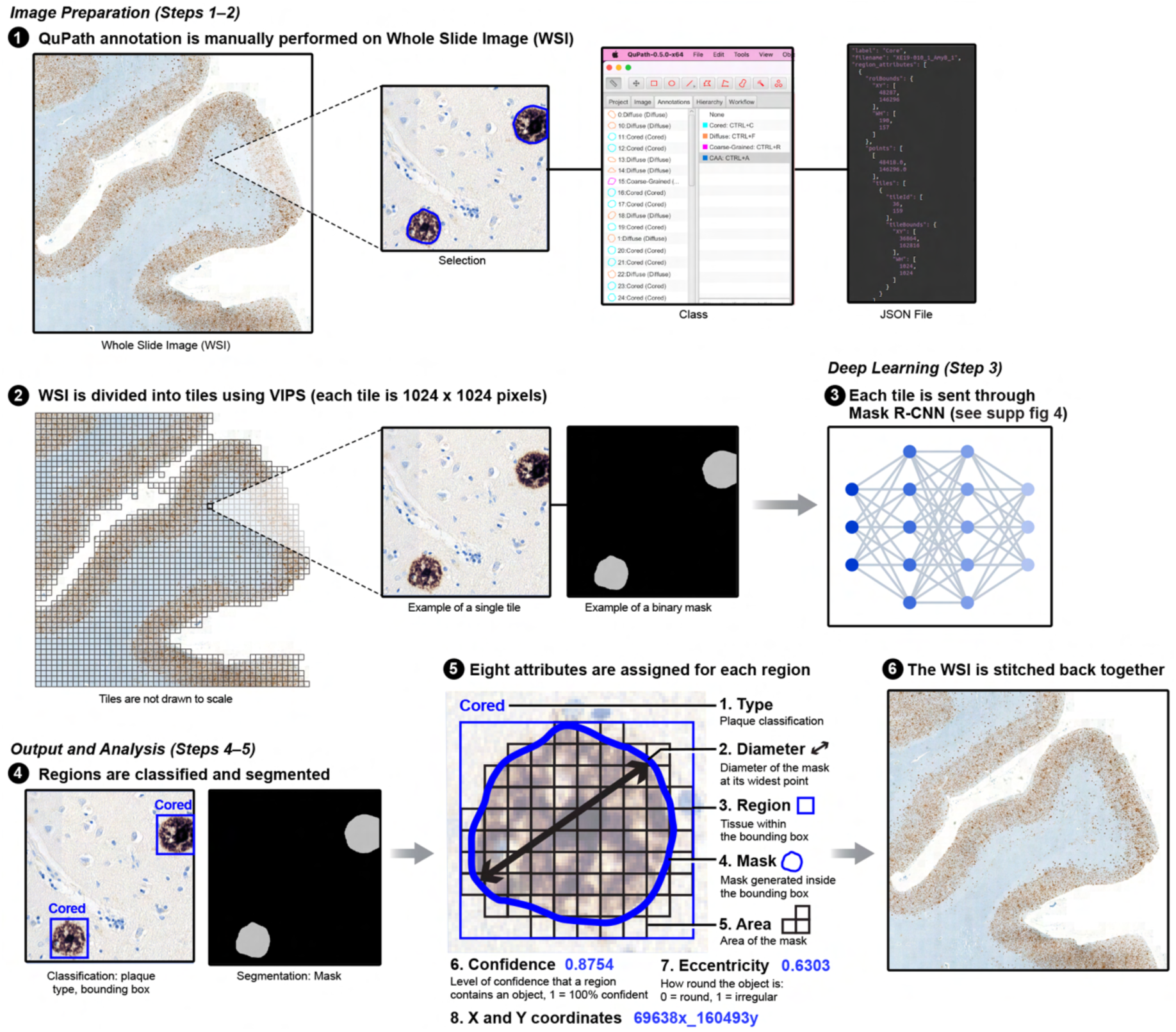
High-throughput and scalable pipeline for digital pathology analysis. 1) Whole Slide Images (WSIs) are annotated using Qupath. 2) The WSIs are broken down to tiles of size 1024 × 1024 for training. 3) The tiles are fed into the Mask Region-based Convolutional Neural Network (Mask R-CNN) along with corresponding binary masks. 4) The model outputs classification and segmentation. 5) Additional attributes are extracted from the segmented masks. 6) The final results are stitched back and visualized at a WSI level.

Existing methods take around 5 minutes per WSI^13^ for classification and 45 minutes per WSI^23^ for segmentation. Our approach, using 1024 × 1024 tile crops, reduces the number of tiles by 16-fold. Combined with processing only the foreground area and implementing the RPN, this reduces the total time to 20 minutes for tile generation, classification, segmentation, and plaque attribute extraction—a 5-fold increase in speed.

### Mask-R-CNN learns to precisely localize, classify and segment Aβ histopathological diagnostic elements on unseen data

To perform supervised DL with a Mask R-CNN, we first needed to develop a training dataset with examples of pathology we wanted it to learn to recognize and examples of nonspecific staining and slide-image level artifacts we wanted it to ignore. Rather than having expert pathologists annotate all the images in the training set, which would be too costly and time consuming, we broke down the annotation process into a series of steps (Fig 2a).

**Figure 2.**
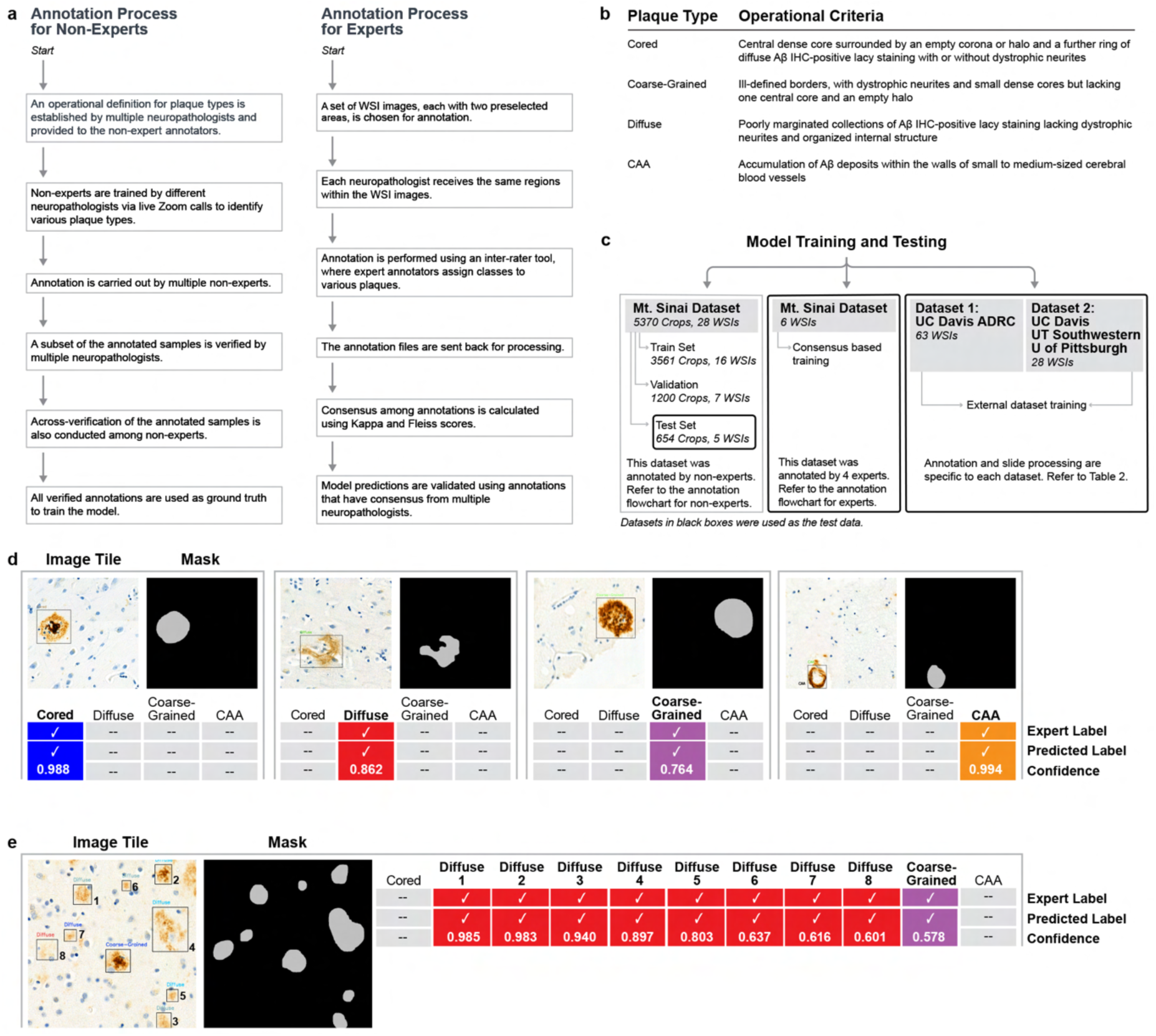
Overview of the annotation process and the model prediction results. a) Annotation process for non-experts (V.G.R., M.A.) and experts (M.S., V.H., B.N.D., O.A.D. b) The operational criteria used for annotation were based on definitions used for research purposes^24, 25^. c) Schematic of the model’s training and testing. d) Model output for the test set obtained from the Mount Sinai dataset. For each tile, both the expert label and the predicted label are shown. Each predicted label also has a confidence score where 1 corresponds to 100% confidence. e) Model prediction on tiles with multiple objects.

First, non-experts (V.G.R., M.A.) learned from expert pathologists (M.S. and V.H.) on how to recognize and annotate four types of amyloid pathologies: cored, coarse-grained, diffuse, and CAA (Fig 2b). Then they manually annotated 23 WSIs consisting of 4716 crops (Supplementary Table 1), submitted a subset of our annotations to the experts (M.S. and V.H.) for verification, and cross-checked our own annotations. We used 16 of these annotated WSIs (3,516 crops) to train the model, and 7 (1,200 crops) as the validation dataset, to check the model’s learning during training. For more details, refer to the ‘Annotation and Dataset Preparation’ section under Methods.

We evaluated Mask R-CNN’s performance using three tests (Fig 2c). First, the model was tested against ground truths established by non-experts trained by experts. Second, it was evaluated against ground truths determined by consensus from 3 out of 4 neuropathology experts. Third, the model was tested on external datasets (Supplementary Table 2): UC Davis (External Dataset 1)^6^, where ground truths were established by individual experts, and UT Southwestern/University of Pittsburgh/UC Davis (External Dataset 2)^9^, where they were based on consensus from 2 out of 4 experts. The results of all these tests are described in this and the following sections.

In the first test we compared the model predictions to the ground truths established on a portion of the original dataset (Mt Sinai Dataset) the model had not seen during training, referred to as the “test dataset”. This dataset comprised 654 random crops from five distinct WSIs and included 946 ground truth annotations. Testing performance this way lessens the chances that model accuracy is misleadingly high because the model has become overfit to features in the training dataset that do not generalize.

Using this test dataset, Mask R-CNN learned to predict both the classification labels and the corresponding segmentation masks for each individual tile with high confidence (Fig. 2d). In addition, the Mask R-CNN was able to classify and segment multiple amyloid pathology objects within a single tile (Fig. 2e), which is important for achieving comprehensive and accurate quantification of neuropathology from the 1024 × 1024 crops.

To ensure that model predictions were compared to ground truth annotations of the same objects, we set the Intersection over Union (IoU) threshold at 0.5^26^. The IoU measures the overlap between an object area predicted by the model and an object area delimited by the annotator. By setting the IoU threshold at 0.5, objects were filtered out that were not outlined properly by the model, which would likely give rise to spurious label and mask predictions. To assess the accuracy of the model, we compared the ground truth labels with the model’s predicted labels and computed “Recall”, the proportion of ground truth plaques detected and correctly labeled by the model, as well as “Precision”, the proportion of model predictions that matched the ground truth annotations. The Dice Coefficient was employed as a performance metric for segmentation: it measures the overlap between the ground truth masks and the model’s output masks. To conserve time and resources, these metrics were computed on a selected subset of crops randomly selected from the foreground tissue area having moderate to frequent plaque densities as annotating all plaques within a given WSI is impractical. Finally, the model’s overall accuracy was determined by comparing all ground truth annotations from the crops within one WSI to the corresponding predictions.

We graphed the precision-recall curve for all object classes across different detection thresholds—defined as the confidence level that determines whether a prediction is valid and is used to classify a detected object as a positive prediction—at a fixed IoU threshold of 0.5. A perfect model would have both precision and recall values of 1 across all detection thresholds. A precision-recall curve is considered optimal when it is positioned towards the top-left corner of the graph. The Mask R-CNN displayed relatively high precision at high recall values (Fig. 3a). To ensure the accuracy assessment is not biased towards a particular class, the F1 score was calculated, which represents the harmonic mean of precision and recall (Fig. 3b), and plotted it against the detection threshold. The F1 scores peaked or plateaued around a detection threshold of 0.6 for all four classes.

**Figure 3.**
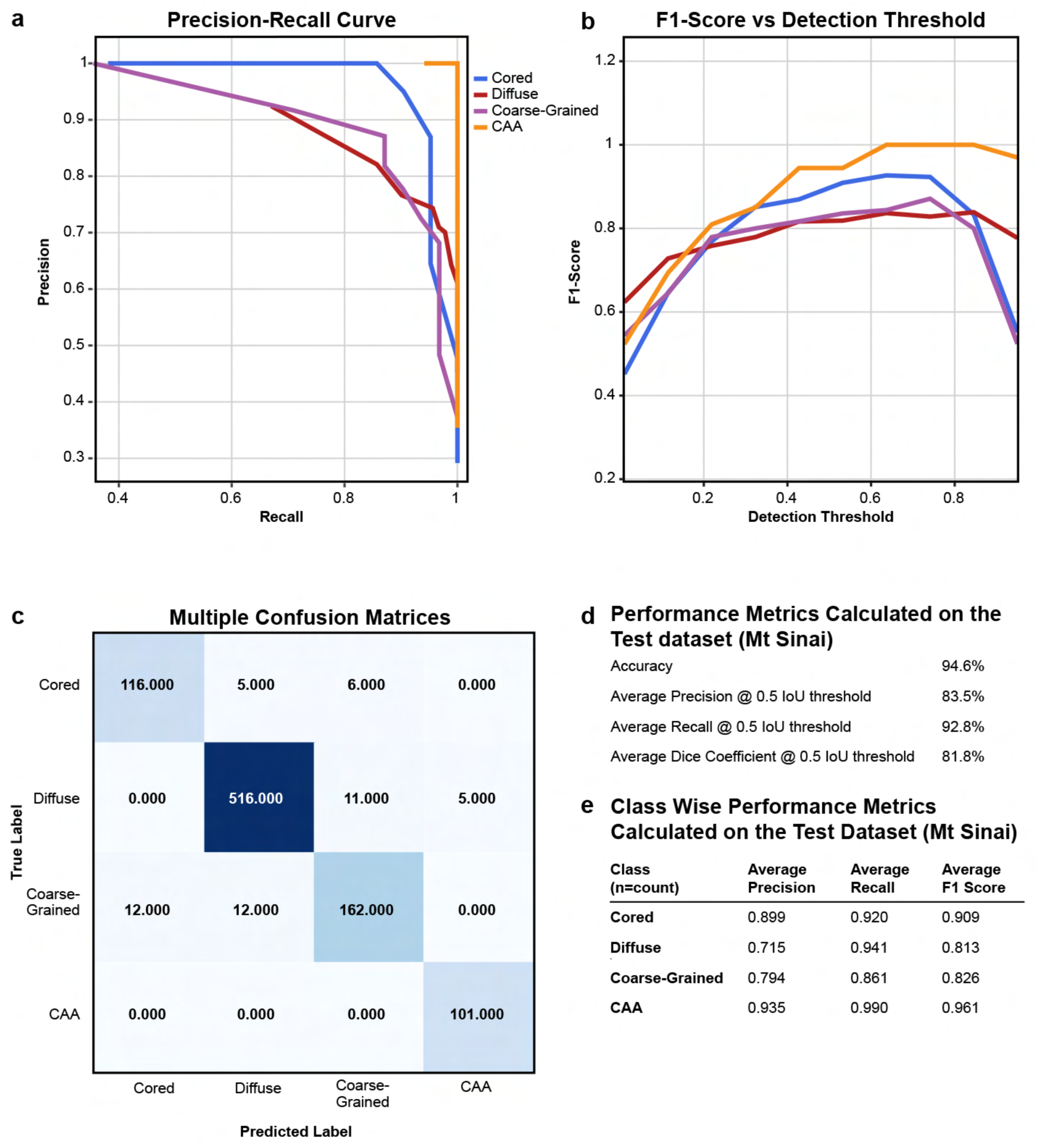
Model performance metrics based on the test set from the Mt Sinai dataset. a) Precision-Recall curve for test obtained for all the four classes - Cored, Diffuse, Coarse-Grained, and CAA - at 0.5 IoU threshold. A curve closer to the top-left corner indicates better model performance. b) F1-scores at different detection thresholds for all the 4 classes. c) Confusion matrix at IoU threshold of 0.5 and detection threshold of 0.6 d) Table shows performance metrics on the test dataset. d) Overall classification performance with precision and recall, and segmentation performance with Dice coefficient, averaged across all classes, at detection threshold 0.6. e) Class-wise classification performance with precision, recall and F1-score at detection threshold 0.6

To visualize the accuracy of the model’s label predictions, we built a confusion matrix, which shows on the diagonal the number of objects assigned the same classification by the model and the ground truth, and off the diagonal the number of objects assigned mismatched classifications (Fig. 3c). In the vast majority of cases, the model’s prediction matched the classification assigned by the ground truth. Indeed, our model showed high overall accuracy (94.6%), as well as high precision (83.5%), recall (92.8%) and F1-score (87.7%) averaged across all the classes (Fig. 3d). An average precision of 83.5% means the model predicted a correct class 83.5% of the time. The overall Dice Coefficient was also high (81.8%), indicating a good overlap between predicted and ground-truth masks, and hence a good segmentation. When examined by class (Fig. 3e), recall varied between 99% (CAA) and 86.1% (Coarse-grained), but precision varied more widely: it was highest for CAA (93.5%) and lowest for Diffuse (71.5%) and Coarse-Grained (79.4%).

### AI Model Shows Stronger Alignment with Consensus Annotations than with Individual Experts’ Annotations

In the second test, to assess the model’s performance against a group of experts, we first needed a benchmark dataset with annotations from multiple experts to establish a consensus-based ground truth. To achieve this, we developed an open source tool in which four experts were asked to label the same set of samples, specifically 2 regions each from six WSIs. We then compared their annotations to measure the level of agreement among them using Cohen’s kappa^27^. In our study, Cohen’s kappa ranged from 0.1 to 0.6, indicating slight to moderate agreement between pairs of experts when labeling the same samples (Fig. 4a). This suggests a fair amount of disagreement among the experts. Additionally, we measured the overall agreement among all four experts using Fleiss’ kappa, which yielded a value of 0.623 (Fig. 4b), indicating moderate agreement across the group.

**Figure 4:**
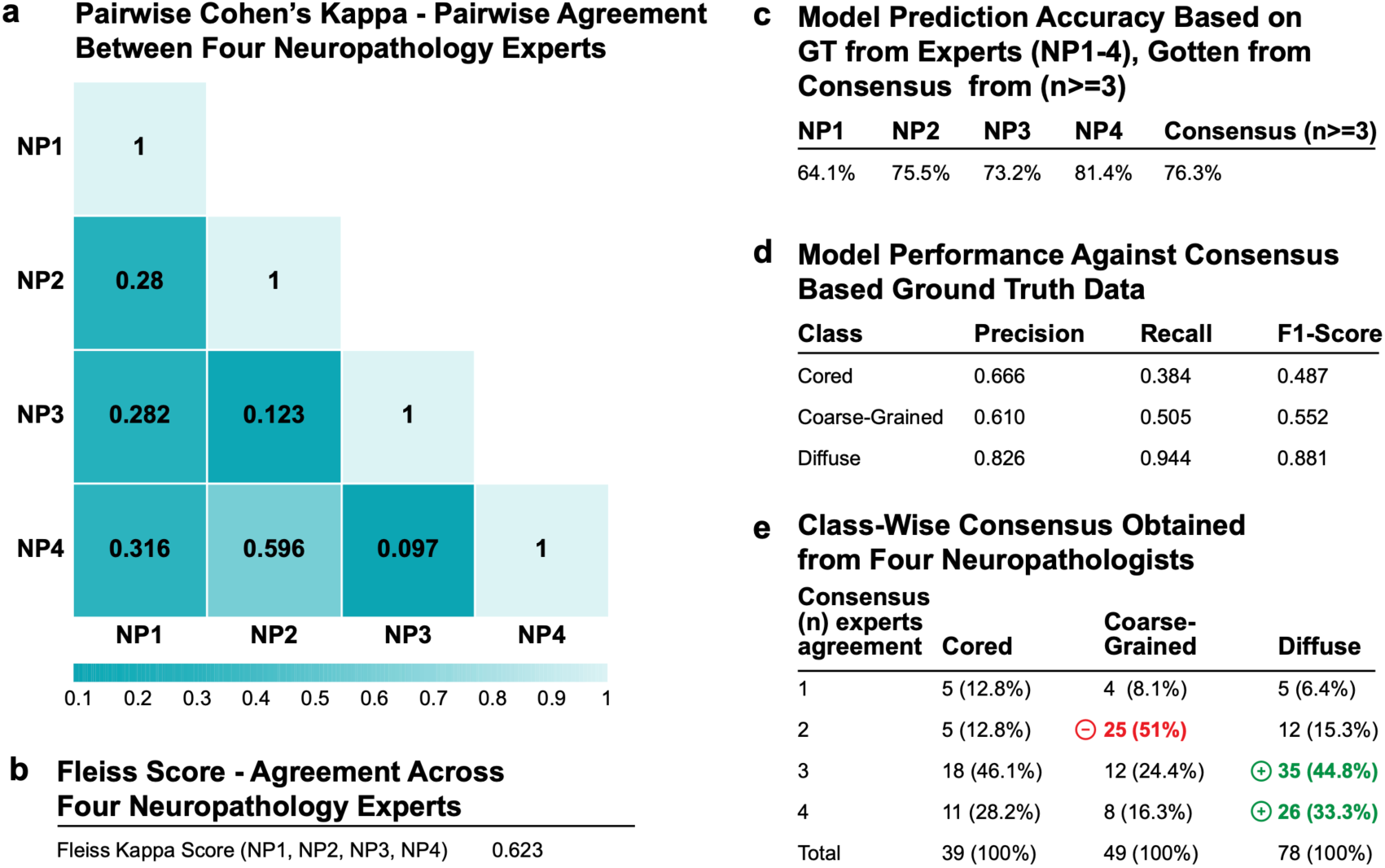
Overview of the consensus agreement between four neuropathology experts. a) Pairwise Cohen’s Kappa agreement between neuropathology experts. b) Fleiss Score agreement between experts. c) Model Prediction results based on the GT from both individual experts and the consensus annotation. d) Model performance against consensus-based GT data. e) Class-Wise consensus data obtained from four neuropathology experts.

We then assessed our model’s accuracy using both the individual experts’ ground truth (GT) annotations or the consensus-based annotations as benchmarks (Fig. 4c). We observed that the model’s accuracy varied between 64.1% and 81.4% depending on the expert’s annotations. This range presumably reflects the disagreement noted above between experts’ annotations of the same data. However, when the model’s predictions were compared to the consensus annotations (n ≥ 3 experts agreeing), the accuracy increased to 76.3%. This was higher than its performance with any individual except for NP4, suggesting the model aligns more closely with the majority or common agreement among experts rather than any single expert’s annotations.

We also found the model’s performance, when tested against the consensus-based ground truth, showed lower precision and recall for certain classes like coarse-grained (Fig. 4d). Upon further investigation of the ground truth annotations from the consensus data, we discovered that there was little agreement among the experts about these classes (Fig. 4e). This suggests that this class is inherently difficult to classify uniformly, possibly because the examples exhibit features of more than one class of Amyloid β plaque, or because experts use different definitions to classify this category.

### Mask R-CNN generalizes well to independent biological datasets for certain plaque classes

To further assess the generalizability of the Mask R-CNN model beyond the Mount Sinai training dataset, we applied it to two independent datasets collected across multiple brain banks. The first dataset consists of 63 WSI AD neuropathology samples from the superior and middle temporal gyri, sourced from the University of California, Davis Alzheimer’s Disease Center Brain Bank^6^. The second dataset^9^ consists of 28 WSIs obtained from three different brain banks: UC Davis (11), UT Southwestern (7), and the University of Pittsburgh (10).

The similarities/differences of features of the datasets including tile size, image magnification, staining reagents, antibodies, brain regions, plaque subtypes (as previous studies did not denote coarse-grained) are described in supplemental Table 2. Given the differences in the datasets and the lack of a standardized benchmarking format—such as file formats and pathology classes (our model is trained on “Cored”, “Coarse-Grained”, “Diffuse” and “CAA”) — we adapted the external datasets for testing in the following ways.

For external dataset 1, the training ground truth data include 256×256 low-resolution images labeled as “Cored,” “Diffuse,” or “CAA.” However, these labels only provided counts and did not include bounding boxes specifying the spatial location of the objects. We reconstructed this information by using the intermediate data from the dataset containing 1536×1536 cropped images and the image_details.csv file that contained the coordinates of both the 256×256 crops and the plaque objects. We then mapped these plaque objects along with their corresponding label to the exact location in the 1536×1536 crops. For generating predictions, we processed the 1536×1536 images by applying a zoom factor of 2, and resizing the crops to 1024×1024. These cropped images were normalized using a reference image from our dataset before being sent to the model for predictions. To evaluate the predictions, we matched the model’s predicted bounding boxes with the ground truth bounding boxes by checking for intersections. The model’s predicted labels were then compared with the ground truth labels for each object.

For external dataset 2, the ground truth images were 1536×1536 in size and included the YOLO bounding boxes with labels. During preprocessing, we applied a zoom factor of 2 to these images, cropped them into 1024×1024 sections, and adjusted the bounding box labels to fit the new dimensions. The 1024×1024 crops were then normalized before being sent to the model for predictions. The ground truth data had two labels: “Cored” and “CAA”. The model performance on this dataset is shown in Table 4b.

**Table 4:**
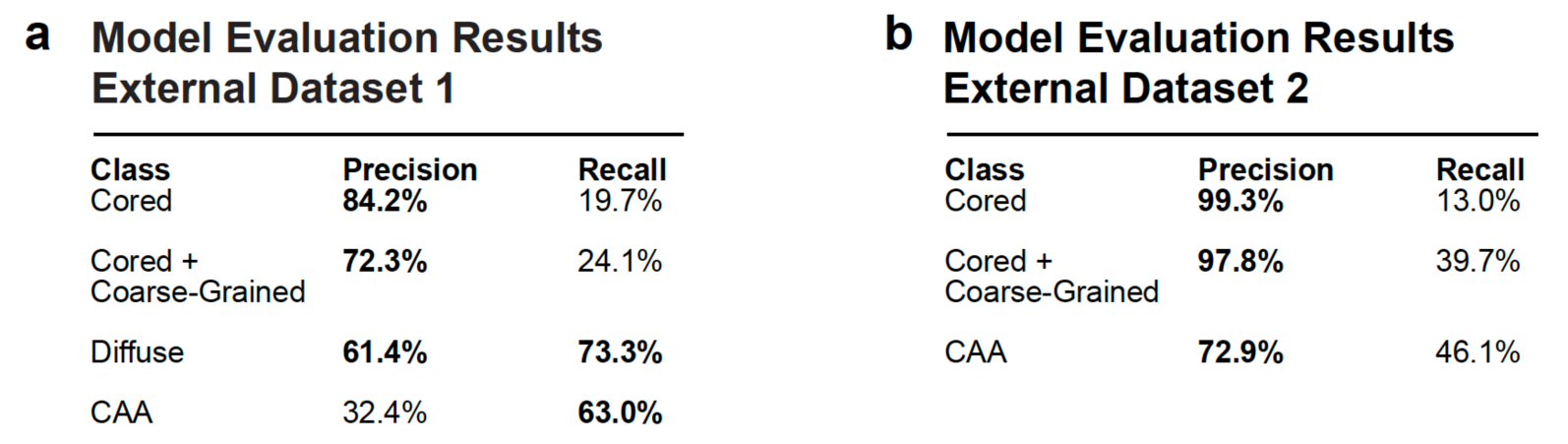
Model prediction results on external datasets. a) Precision-Recall metrics obtained for model prediction on external dataset 1. b) Precision-Recall metrics obtained for model prediction on external dataset 2.

| a Model Evaluation Results<br>External Dataset 1 |  |  | b Model Evaluation Results<br>External Dataset 2 |  |  |
| --- | --- | --- | --- | --- | --- |
| Class | Precision | Recall | Class | Precision | Recall |
| Cored | 84.2% | 19.7% | Cored | 99.3% | 13.0% |
| Cored +<br>Coarse-Grained | 72.3% | 24.1% | Cored +<br>Coarse-Grained | 97.8% | 39.7% |
| Diffuse | 61.4% | 73.3% | CAA | 72.9% | 46.1% |
| CAA | 32.4% | 63.0% |  |  |  |

When we compared the ground truth and model predictions for “Cored” plaques, we noticed that many “Coarse-Grained” plaques in the model’s predictions were labeled as “Cored” in the ground truth data coming from both the external datasets(see Supplementary Figure 6). This discrepancy affects the model performance. To make the class definitions uniform, we merged the “Coarse-Grained” plaques as part of the “Cored” plaques which improved our recall for “Cored” objects to 24.1% and 39.7% (Table 4 a & b, row, Cored + Coarse-Grained)

Our model demonstrated good performance for all the classes on the external dataset,despite variations in tile size, biological sample preparation, and labeling between the training and testing datasets (Supplementary Table 2). However, the recall performance on the “Cored” class was modest across both datasets, a recall of 19.7%, and recall of 13% in both external dataset 1 and 2. This is due to the inconsistent definitions of “Cored” between datasets (Supplementary Figure 5) and the introduction of a new class, “Coarse-Grained” in our own annotation of the training dataset. Overall, the model’s generalization is influenced by differences in input image characteristics and labeling conventions, which affect performance consistency across classes and datasets.

### Mask R-CNN focuses on features inside the pathologies to make its classification

The relationships that form between the layers of trained models such as our Mask R-CNN are non-linear and too complex to interpret directly; as a result, it is difficult to determine how the trained models make the classifications they learn to make. New tools such as Ablation-CAM^28^ have been developed to reveal to the human eye features in the image that the network learned to use to perform classifications, and so we used it here to understand what important features the model uses to predict the different types of plaques.

To implement Ablation-CAM, we first employed a gradient-free localization approach to obtain the feature maps. Then we ablated the feature maps randomly, and visualized the quality and accuracy of pixel-wise predictions. After repeating these steps on all the feature maps, we obtained a heatmap of pixel areas that the Mask R-CNN learned to use to make its predictions Supplementary Fig. 2). We found the pixels the Mask R-CNN learned to use to correctly classify each pathology were primarily and consistently within the pathology object itself. These may include variations in the intensity and spatial distribution patterns of positive pixels within the pathologies as reliable indicators of class, not unlike the approach of an expert.

### Predictions from Mask R-CNN overlap with the CERAD ratings

CERAD has established standardized procedures for the evaluation and diagnosis of persons with AD^29^, including some of the best-validated tools to relate amyloid pathology to disease severity^3^. The CERAD ratings are calculated by evaluating the density of neuritic plaques in specific brain regions, categorizing them as none, sparse, moderate, or frequent based on a semi-quantitative estimate by experts. We wondered if the automated measures of amyloid pathology made by the Mask R-CNN from postmortem brain samples correlated with the clinical features such as CERAD ratings of the severity of AD.

To investigate, we applied our trained model to identify and quantify plaques in a test dataset comprising 298 WSIs. The plaque quantification was compared with the CERAD ratings calculated independently by trained pathologists (D.P, V.H) sometimes a decade before the initiation of the current study in the Middle Frontal Gyrus region of the brain. The median value of Cored plaques per whole slide image is 429 for “frequent”, 124 for “moderate”, and 15 for “sparse” rating. Our model-predicted plaque counts aligned very well with these ratings (Fig. 5a). We also observed a stark contrast between the high dynamic range ratio-scale counts generated by the model and the low dynamic range ordinal scale gleaned from CERAD. This underscores how classification as sparse, moderate, and frequent misses the likely biological differences contained within these restrictive classifications.

**Figure 5.**
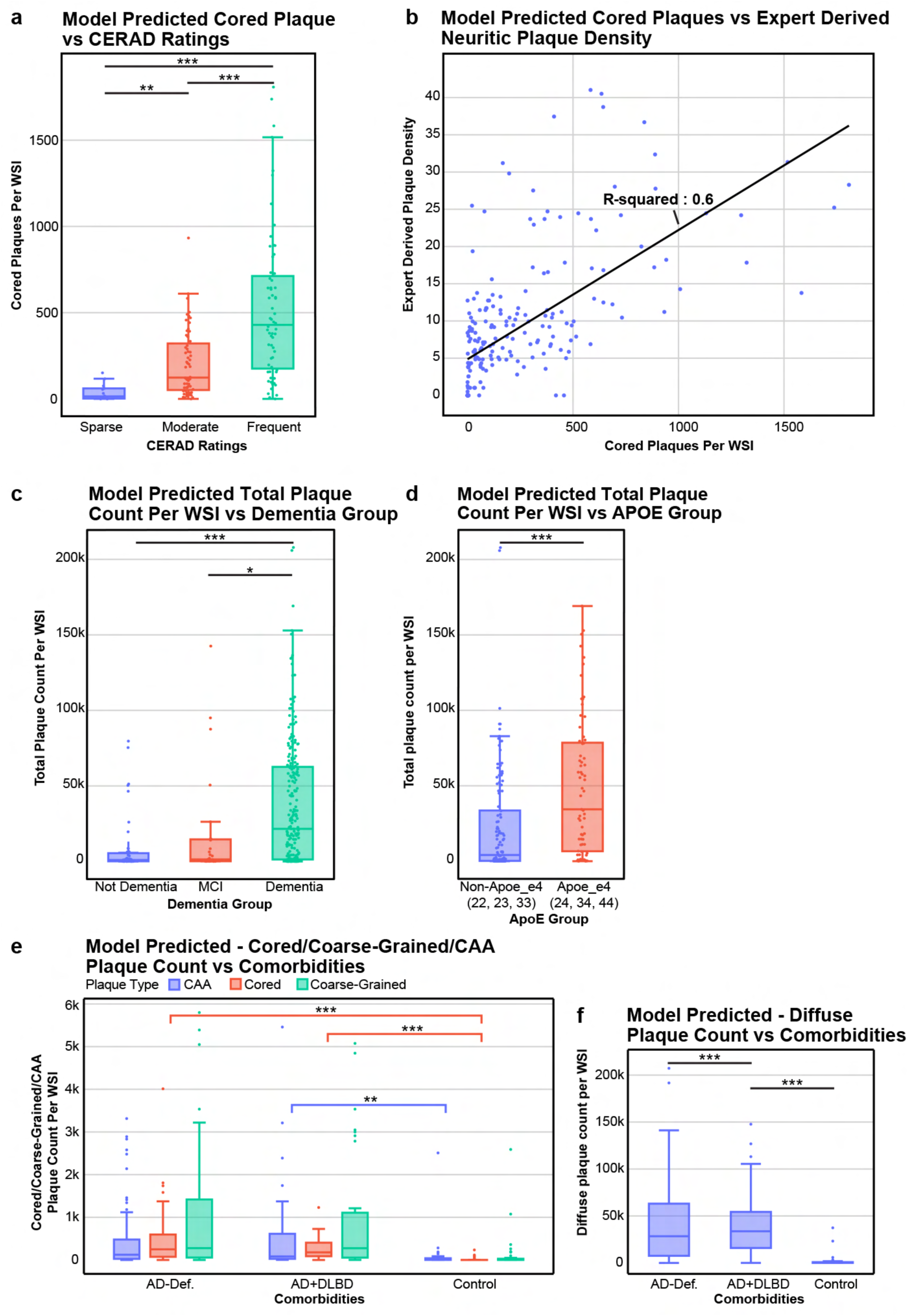
Correlation of model’s predicted plaque counts with expert-derived CERAD ratings and APOE groups. a) Predictions from Mask R-CNN overlap with the CERAD ratings. Box plot showing model predicted total neuritic plaques with cores vs the expert based CERAD ratings: sparse, moderate, and frequent. One-way ANOVA test gives F-val 22.6 & p-val 5.1e-9; Asterisks indicates significance between two groups - “*” for p-val<0.01, “**” for p-val<0.05 and “***” for p-val<0.001 b). Predictions from Mask R-CNN correlates with average density of neuritic plaques for each person. Scatter plot showing model-predicted neuritic plaques with cores on x-axis, and average density of neuritic plaques on y-axis. A regression line is fit having an R-squared value of 0.6 c). Plaque counts reveal key differences across dementia stages. Box plots showing correlation of total plaque counts per WSI vs Dementia groups. ANOVA test shows that the three groups are significantly different with F-statistics - 13.7, p< 1.9e-6. d). Plaque counts reveal key differences across APOE-e4 status. Box plots showing correlation of total plaque counts per WSI vs ApoE groups. Independent t-test shows significant difference with p<2.03e-5. e) Plaque counts reveal significant differences with control. Box plots showing difference of total plaques between AD Def and Control with independent t-test (p-val < 2.57e-9), and AD + DLB and Control with independent t-test (p-val < 1.57e-11)

To validate the correlation among the different frequency groups—“sparse,” “moderate,” and “frequent”—we conducted a one-way ANOVA test. The results revealed a highly significant difference between the means of the groups, with an F-statistic of 22.06 and a p-value of 5.1e-9. To further explore these differences, we performed independent t-tests, which allow for pairwise comparisons between the groups. The results showed a very highly significant difference between the “frequent” and “moderate” groups (p < 6.16e-7), as well as between the “frequent” and “sparse” groups (p < 7.08e-5). Additionally, we found a highly significant difference between the “moderate” and “sparse” groups (p < 0.0017).

We also found a positive linear correlation between the total number of neuritic plaques with cores predicted by the model and the expert-derived average density of neuritic plaques, calculated as the number per mm² across five regions per slide(Fig. 5b)^30^. Furthermore, a Spearman Correlation performed between expert-based average neuritic density and model-predicted cored plaques per WSI yielded a statistically significant coefficient value of 0.764 (p≤ 2.7e-44).

### Plaque Counts Reveal Key Differences Across Dementia Stages and APOE-e4 Status and Comorbid Conditions

We also found a significant correlation between the “No Dementia” (N=51), “Mild Cognitive Impairment” (N=27), and “Dementia” (N=220) groups and the total number of cored plaques per WSI. The Dementia status was based on the donors cognitive state during the non-agonal 90 days prior to death^30^. By ANOVA test, the three groups were significantly different (F-statistics - 13.7, p≤ 1.9e-6) Pairwise comparisons were made with an independent t-test, and they revealed differences between “Dementia” and “No Dementia” group (p≤ 1.8e-6) and between “Dementia” and “MCI” (p≤ 0.018) but no significant difference between “MCI” and “No Dementia” (p ∼0.13)(Fig. 5c).

Next, we investigated whether plaque count was correlated to APOE - e4 status by comparing individuals with APOE allele genotypes of e2/e4, e3/e4, and e4/e4 to those with APOE genotypes of e2/e2, e2/e3, e3/e3, etc. With an independent t-test, we observed a significant difference between plaque count in persons with at least one e4 (N=69) allele versus those with no e4 alleles (N=120) (p ≤ 2.03e-5) (Fig. 5d).

Furthermore, we observed a high count of amyloid pathologies in persons identified as “Definite AD” cases (AD Def, N=146) and “AD & Diffuse Lewy body disease” (AD+DLBD, N=42) as compared to persons labeled as “Control” (N=47) (Fig. 5 e,f). There is no significant difference in plaque count between the two groups AD Def and AD+DLBD which means plaque counts remain high regardless of Lewy body disease. Independent t-tests conducted between the comorbidity groups, AD Def. and Control (p-val < 2.57e-9), and AD+DLBD and Control (p-val < 1.57e-11), revealed significant differences in plaque count compared to the Control group.

## Discussion

In this study, we developed an open-source tool for collecting consensus annotations from both experts and non-experts, as well as a scalable computational pipeline to process large volumes of whole-slide images (WSIs) for comprehensive, unbiased, and quantitative neuropathology assessments. Using this tool, we established a ground truth dataset for training and validation. The dataset was then used to train a model capable of accurately classifying and segmenting various types of amyloid pathology—cored plaques, coarse-grained plaques, diffuse plaques, and amyloid angiopathy—with human-level precision. To ensure reproducibility, we packaged this tool as a plugin for QuPath, allowing researchers to obtain prediction results without requiring dependency installations or dedicated GPU infrastructure.

Several advances in the design of the computational pipeline contributed to its success. First, leveraging GPU architecture(Nvidia RTX A4000) and using 1024 × 1024 image crops instead of the more widely used 256 x 256 crops improved performance in two key ways. Based on feedback from expert neuropathologists, the larger crops provided critically needed context for accurately annotating images in the training dataset. Additionally, the larger crops led to a ∼5-fold increase in throughput, reducing processing time to just 20 minutes per slide. Second key advance was the use of Mask R-CNN architecture. Incorporating the Region Proposal Network (RPN) improved the efficiency of the pipeline by focusing in-depth analysis on regions with a high probability of containing neuropathological features. This architecture also allowed for simultaneous identification and segmentation of neuropathological objects, enabling not only counts but also deeper analysis of features such as size, shape, and texture.Although this capability was not fully utilized in this study, we anticipate future work could reveal correlations between these sub-features of pathology and specific patient donor characteristics and relationships of the different neuropathological features to different molecular or proteomic signatures.The third key advance was the development of an API (Application Programming Interface) along with a QuPath plugin for running model inference through a user-friendly GUI interface, with the model deployed on a GPU cloud.

Several observations suggest our model emulates the human learning and annotation process. First, we found a significant, positive correlation between independent expert-estimated amyloid β pathology burden, as measured by CERAD plaque density scores, and the model’s automated amyloid pathology measurements. This automated quantification in postmortem brain tissue also significantly correlated with AD severity and linked higher numbers of cored plaques to both dementia severity and APOE e4 status, replicating findings from other groups ^8,31,32,33,34^ on independent datasets and supporting the model’s reliability. An important future direction will be to leverage the capability of the Mask R-CNN to segment pathological objects to determine whether more specific features of certain types of amyloid pathology have an even stronger predictive value for AD severity or specific symptoms of persons with AD. Second, our Ablation-CAM experiment shows the Mask R-CNN model bases its predictions on pixel patterns within pathological objects, similar to trained human annotators (Supplementary Fig. 2), giving us confidence that it learned essential features of amyloid pathologies. Third, when tested on datasets from two independent brain banks, the model achieved high prediction accuracy for classes like diffuse plaques and amyloid angiopathy (CAA), demonstrating its ability to generalize amyloid pathology features.

Few published models have been tested beyond the dataset on which they were trained^15^, in part because it has been difficult to obtain publicly available annotated AD neuropathology datasets. Unfortunately, without testing algorithms on independent datasets, it is difficult to denote generalizability. This is a concern with digital pathology because of the many potential ways in which isolated training datasets may have unique features that could be learned by DL networks and would limit their generalizability^35,10^. For example, different brain banks often have slightly different sample preparation techniques, and the intensity of labeling can vary from sample to sample, even within the same brain bank^36^. Since we tested the generalizability of our algorithms on datasets generated by independent brain banks on an independent cohort of persons who donated their brains, all the potential issues described above could be relevant. In addition, the independent dataset came in the form of 256 x 256 image crops. To be able to assess the performance of our algorithms on those data, we first needed to resize the crops to 1024 × 1024, an operation that almost certainly caused some loss of detail and sharpness in the resultant images. Nevertheless, our models were able to predict the three types of amyloid pathology in these resized images. Unsurprisingly, the performance of the models was not as high on this new dataset as it was on the validation set from Mt Sinai, underscoring the importance of standardized annotations, consistent image formats, and establishing benchmarks for model generalizability.

The accuracy of the supervised machine learning approaches used here is inherently limited by the accuracy and reproducibility of the human annotation used to generate the dataset to train the model. Algorithms may learn from annotations to accurately mimic the predictions of one expert but their accuracy would fall if they were applied to a different dataset annotated by an expert with different judgments.

For example, the lower recall for classes like cored could be because of slightly different definitions used for annotating the two external datasets and the limited availability of consensus data for this class for training and evaluation. To address this limitation, we developed tools to collect consensus annotations from multiple experts. Indeed, our inter-rater study of 4 experts annotating the same dataset found that their disagreement was greatest on their classification of coarse-grained objects. We also found the model demonstrated better performance when evaluated against consensus annotations (n ≥ 3 experts agreeing) compared to individual expert assessments. This suggests consensus-based data provides a stronger, more consistent ground truth for training and evaluating the model, particularly in cases where individual experts may have differing interpretations. This finding supports the use of consensus annotations to enhance the generalizability of the model’s ability to perform complex classifications.

The trained models were then used to determine whether and to what extent the quantification of amyloid β pathology in postmortem material from deceased individuals correlates with the severity of AD neuropathology as measured by CERAD scores, or severity of dementia proximal to the end of life. We found there was a positive and significant correlation between the expert estimated plaque burden as well as dementia severity and the unbiased automated quantitative measures of amyloid pathology. An important future direction will be to leverage the capability of the Mask R-CNN to segment pathological objects to determine whether more specific features of certain types of amyloid pathology have an even stronger predictive value for AD severity or specific symptoms and neurobiological features of the AD patients.

We acknowledge a few caveats in this study. First, the inter-rater annotation study was conducted on a single brain region (Middle Frontal Gyrus), stained with AB 4G8 antibody from Mt. Sinai dataset with limited WSIs (6). Future studies should include multiple brain regions, stained with different antibodies, from various brain banks to fully realize the benefits. Currently, the training dataset was annotated by non-experts who learned from the inter-rater study results. In the future, the development of large training datasets based on consensus annotations might further improve the generalizability of the models trained from them. We aim to develop training datasets annotated exclusively by a team of experts and also fine tune the existing model with these updated annotations. Additionally, quantifying and segmenting diffuse plaques remains inherently challenging due to their poorly defined structures and boundaries. However such quantification may prove especially important in cases of presymptomatic or incipient AD ^37^. Importantly, neuropathology evolves as disease progresses, and so it is quite likely that some examples of pathology may represent intermediates between two more well-defined classes with some features of each class. Such edge cases pose an inherent challenge to the human annotator as well as to the AI model, and are likely a major contributor to the disagreements amongst annotators in our interrater study and between annotators and the AI model. One major advantage of the use of AI models is that they make the same prediction consistently and reproducibly, increasing the sensitivity of comparisons made between subjects.

In sum, this work highlights the advanced capabilities of our pipeline in analyzing large sets of WSIs. We developed algorithms that identify and classify various types of amyloid β pathology with human-level accuracy. These algorithms not only generalize well to independent datasets but also provide measures of amyloid β pathology that correlate strongly with the widely accepted CERAD score and analyses with these algorithms replicated other findings including an increase in amyloid pathology in brain tissue from persons diagnosed with AD with an APOE epsilon 4 allele^34,8^. This robust analysis capability enables large-scale explorations of correlations between neuropathological features of AD and diverse data, including clinical features and genotypes. Such insights could reveal how genetic variants contribute to different types of central nervous system pathology through both cell-specific effects and interactions between various cell types and hallmark pathology. The analysis is integrated into a user-friendly QuPath interface for broad clinical and research use.

## Methods

### Ethics approval

These studies utilized tissue obtained from postmortem human brain specimens. All data followed current laws, regulations, and IRB guidelines (such as sharing de-identified data that does not contain information used to establish the identity of individual deceased subjects). De-identified data do not contain personal health information (PHI) like names, social security numbers, addresses, and phone numbers.

### Case Cohort and Sample Preparation

The Mt. Sinai dataset used here includes a total of 298 cases obtained from Mount Sinai Brain Bank, all of which are diagnosed with Alzheimer disease. A medical record review was performed on all cases using a structured instrument that records and codes all medical and neuropsychiatric conditions (based on ICD-9/10 and DSM IV/V codes), all medications (including doses and duration), the severity of dementia (based on clinical dementia rating), neuropsychiatric symptoms (NPS), family history, available laboratory and neuroimaging study results and sociodemographic variables such as level of education and languages. Brain sections from middle frontal gyrus were prepared from each AD case in paraffin-embedded blocks and immunolabeled with anti-β amyloid (4G8). Neuropathological diagnostic criteria for AD are based on CERAD, which includes quantification of neuritic plaques (NP) in the middle frontal gyrus brain region. These WSI were a subset of WSIs that are publicly available at https://mountsinaicharcot.org/. External dataset 1 (UC Davis)^6^ and external dataset 2 (UC Davis, UT Southwestern and University of Pittsburgh)^9^ were downloaded from https://zenodo.org/records/1470797 and https://zenodo.org/records/7944799 respectively.

### Pipeline

#### Inter-Rater Tool Development

The inter-rater tool is developed as a QuPath Plugin (version 5.0) and is licensed as open-source software. The tool can be used by anyone to conduct inter-rater variability studies. Before conducting the inter-rater study, the researcher must add WSIs to a project. The tool allows researchers to navigate to an object to annotate and enables zoom-in and zoom-out to assess the object’s surroundings. Depending on resources and time constraints, 1 or 2 squares 1mm x 1mm are selected per WSI. In our case, we selected 2 regions across 6 WSIs that covered all types of classes and their variations so that our model could learn all the classes and the edge cases correctly. This entire process takes around 1-5 minutes per WSI depending upon the skill level of the annotators. After this step, the project files are compressed and sent to all the other raters. Each rater will have the exact same regions to annotate (Fig. 6, Step 1) To make annotations faster and easier, we include a set of hotkeys depending on the type of annotation done. For amyloid β plaques, we use Ctrl+C for core, Ctrl+F for diffuse, and Ctrl+A for CAA, as shown in (Fig. 6, Step 2). This configuration can be changed for other applications. After annotations are done, the tool automatically generates a JSON file containing all the annotations. In (Fig. 6, Step 3), the JSON files from different raters are collected and processed using a Python script. We calculate Cohen’s kappa for pairwise comparison between two raters and Fleiss’ kappa for comparison across all raters. (Fig. 6, Step 4) is optional; it overlays all the annotations from different raters the image regions to understand the variability in annotations. A video demonstrating annotation with the tool can be accessed [here](https://www.dropbox.com/scl/fi/2ssy1mq3ju7ph82hfciv7/Interrater_tool_demo.mp4?rlkey=ut8sqf1c5e7kvj15op7qn4pgv&dl=0).

**Figure 6.**
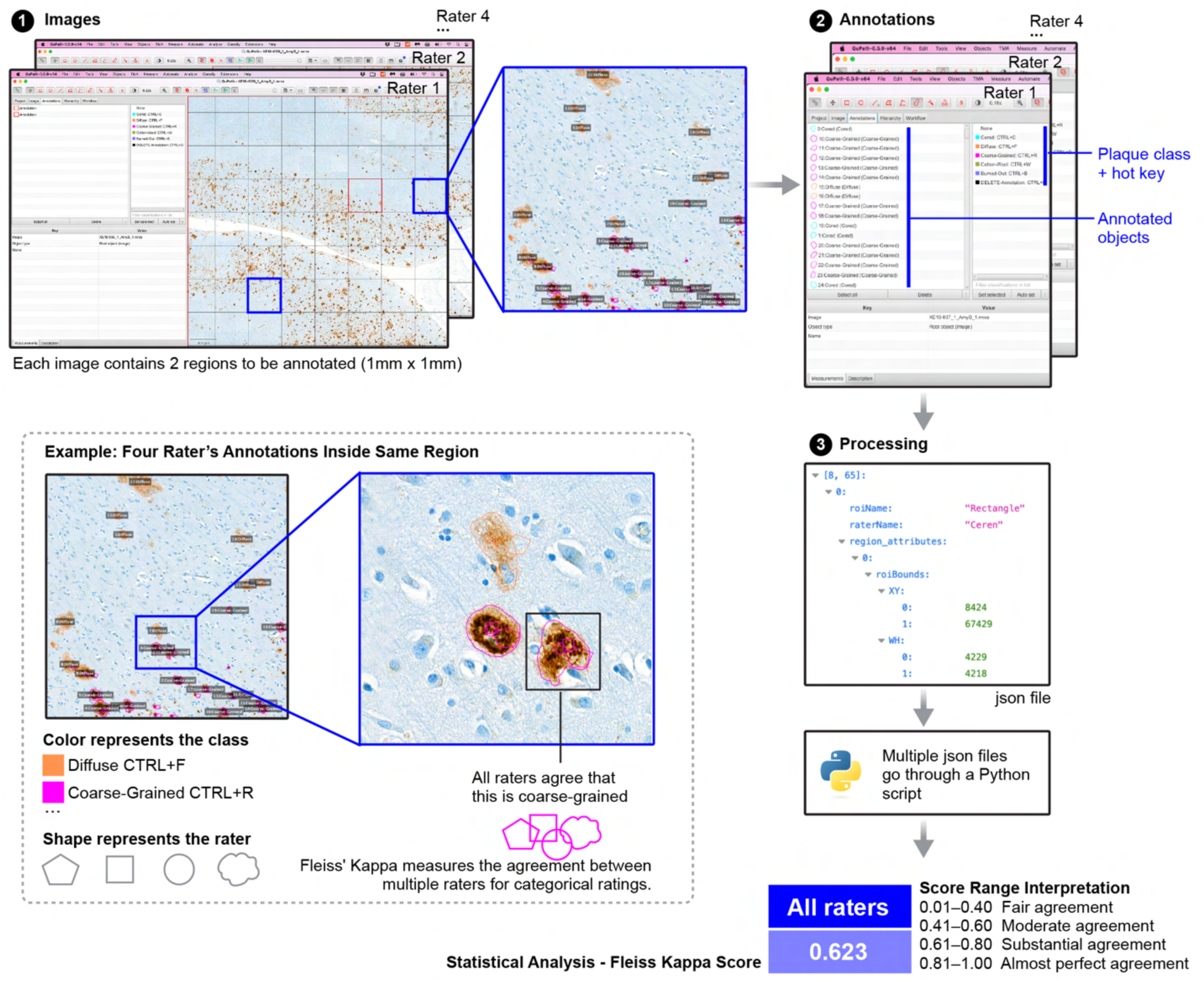
Inter-rater tool as a plugin in Qupath. Researchers add WSIs to a project, select 1mm x 1mm regions, and share compressed project files with raters. Hotkeys simplify annotation (e.g., Ctrl+C for core plaques). After annotations, a JSON file is generated. The data is processed to calculate Cohen’s kappa for pairwise and Fleiss’ kappa for overall comparison. Optionally, annotations are overlaid to visualize variability.

#### Annotations and Data Preparation

The annotation of the training set was done by non-experts (Fig. 2, left column). First, experts provided an operational definition (Fig. 2b) to standardize the approach among all annotators. Second, annotators reviewed samples annotated in an inter-rater study to refine their understanding of the object to annotate. Then, the annotators annotated the training dataset. The training set WSIs were loaded into QuPath at 20X magnification. The resolution was set to 1 micron per pixel (MPP) and a 1024 × 1024-pixel grid was added to the Image View. The annotators annotated the grids that contained Cored plaques, Coarse Grained plaques, Diffuse plaques or CAA. Pixel-level segmentation of the plaque regions was done with a magic wand/polygon tool. The annotations were written into a custom JSON format containing all the important polygon regions and their corresponding labels.

The Mt. Sinai training dataset consisted of 5,370 crops of size 1024 × 1024 pixels obtained from 28 WSIs and included classification labels and pixel-level segmentation masks. To provide more variation in the training cycle, objects in the dataset were not centered within the crops and the dataset was augmented using various techniques (Supplementary Table 3) from the Albumentation library^38^. The dataset was divided into a training dataset consisting of 3516 crops extracted from 16 WSIs, a validation dataset containing 1,200 crops obtained from 7 WSIs, and a test dataset containing 654 crops extracted from 5 WSIs (Figure 2c, left box).

#### Model Development and Training

The Pytorch framework was used for the development of the model. The architecture is based on the Mask Regional Convolutional Neural Network (Mask R-CNN)^22^. The Mask R-CNN model is split up into three stages. The first stage uses a Feature Pyramid Network (FPN)^39^ to extract the features from an input image at different scales (Supplementary Fig. 4). A 1 x 1 convolution filter is applied to all the feature maps from the corresponding image scales and the resulting feature maps are averaged to form the final feature map. This step ensures that the feature map contains important information from both low resolution and high resolution images for a given object of interest. We use the pre-trained weights from ResNet50 as our backbone architecture for the FPN model.

The second stage (Supplementary Fig. 4) includes a lightweight neural network called the Region Proposal Network (RPN) that scans the feature map generated by an FPN to propose regions that may contain a plaque. RPN relies on anchor points and anchor boxes. Every point in a feature map is an anchor point. We generate anchor boxes for every anchor point. The anchor boxes help us to map the features from the feature map to the raw image location in the input image. A set of 9 anchor boxes are generated for each anchor point based on different combinations of scale and aspect ratios.

To get good quality anchors, a Non Maximal suppression algorithm is used where all the anchors whose intersection of union (IoU) with the ground truth (GT) box is below 0.7 are discarded. The RPN takes these anchors and uses a fully connected layer to give two outputs: (1) - binary class - whether an anchor contains an object or not and (2) the bounding box regressor representing the delta in x, y, w, and h that are required to come closer to the GT boxes.

In the third stage (Supplementary Fig. 4), the network takes in the regions proposed by RPN as input and uses Region of Interest (ROI) align pooling to extract the corresponding feature map without quantization. This feature map is sent to a set of CNN’s for generating pixel-wise classification and segmentation masks. We use the following loss function:

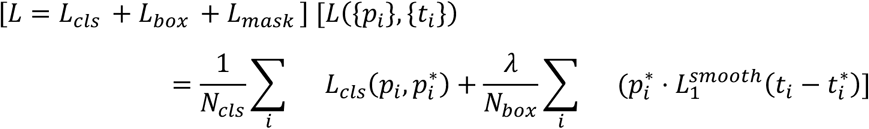

where *L*_*cls*_ is the log loss function over two classes. For the box loss, we can easily translate a multi-class classification into a binary classification by predicting a sample being a target object versus not as 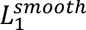 loss.

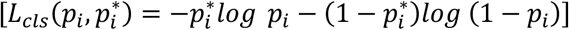

*L_mask_* is defined as the average binary cross-entropy loss, only including *k* the mask if the region is associated with the ground truth class *k*.

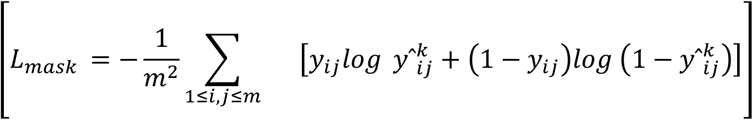

where *yij* is the label of a cell (*i*, *j*) in the true mask for the region of size mxm; 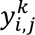 is the predicted value of the same cell in the mask learned for the ground-truth class *k*.In this phase, the model is fed only the crops annotated by experts containing objects of interest. The training dataset described in the data preparation step is used for training the model, and the validation dataset is used by the model to evaluate its performance on every epoch. The Mask R-CNN model is trained on four NVIDIA A40 GPUs with pytorch lightning with the ResNet152 pretrained model as backbone. The model is trained for 150 epochs with a batch size of 8. We use AdamStochastic Gradient Descent as the optimizer of the model with a learning rate of 10^-43^ and a weight decay of 10^-6^. The training loss and validation metrics such as loss, classification accuracy, segmentation dice coefficient are monitored for each epoch to avoid overfitting and to ensure that the model is learning properly. The best model is saved as learned parameters optimized on the highest validation accuracy.

#### Model Testing, Inference and Explaining Model Predictions

The model’s performance is evaluated using three different tests (Fig. 2c). Once tested and deployed, we apply the model to the entire Mt. Sinai Dataset to infer and quantify plaques. During this phase, the model processes only the tissue regions of the WSI, identified by downscaling the image by a factor of 4 and applying Otsu thresholding to generate a contour mask (Supplementary Fig. 3). This step allows us to extract tile coordinates of the tissue areas, which are saved as a numpy file of approximately 100 KB.

For inference, we utilize an NVIDIA RTX A4000. The pipeline takes the whole slide image and its corresponding numpy file to extract the relevant crops and run inferences with our trained model. The model detects plaques, predicts their labels, and generates segmentation masks (Fig. 2d). Additionally, it extracts features such as area, diameter, and brown pixel count for each segmented object.

The inference pipeline is optimized for multiprocessing, enabling processing of each WSI in about 20 minutes, although exact timing may vary depending on the hardware used. Finally, the model predictions are linked with clinical metadata (Fig. 5).

To understand what features the model uses to predict the different types of plaques, we use Ablation-CAM^28^. In this technique, we employ a gradient-free localization approach to visualize the predictions from the model (Supplementary Fig. 5). The feature maps generated from the FPN model are randomly ablated to check how they affect the pixel-wise predictions. y^c^ is the probability of class C. y^ck^ is the probability of class C after ablation. If a certain pixel is important, then the difference between y^c^ and y^ck^ will be large. This is done repeatedly on all the feature maps. The final output of Ablation-CAM is a heatmap of pixel areas that are critically important for the prediction.

#### AI Model Deployment for Plaque Detection in Whole Slide Images

Model deployment and inference are critical processes in the machine learning lifecycle. Inference is the process by which a model generates predictions on real-world test data. To facilitate use by researchers and clinicians who may not have extensive computational expertise, we developed an open-source tool with a user-friendly graphical interface. This tool allows researchers to upload whole slide images (WSIs) and generate predictions like plaque localization and detections, counts, and segmented binary masks. The tool is developed as a QuPath script. To get started, the user downloads the script and opens it up in the QuPath script editor. Then the user uploads a WSI containing amyloid beta plaques and runs the script. Next the user selects a region of interests by drawing a square box with a defined size of 1024 × 1024 pixels. After the “run inference” button is pressed, the results are generated and overlaid on to the WSI within 5 seconds. All the additional attributes (e.g., plaque count, area, eccentricity, etc…) can be seen on the script editor terminal.

**Figure 7.**
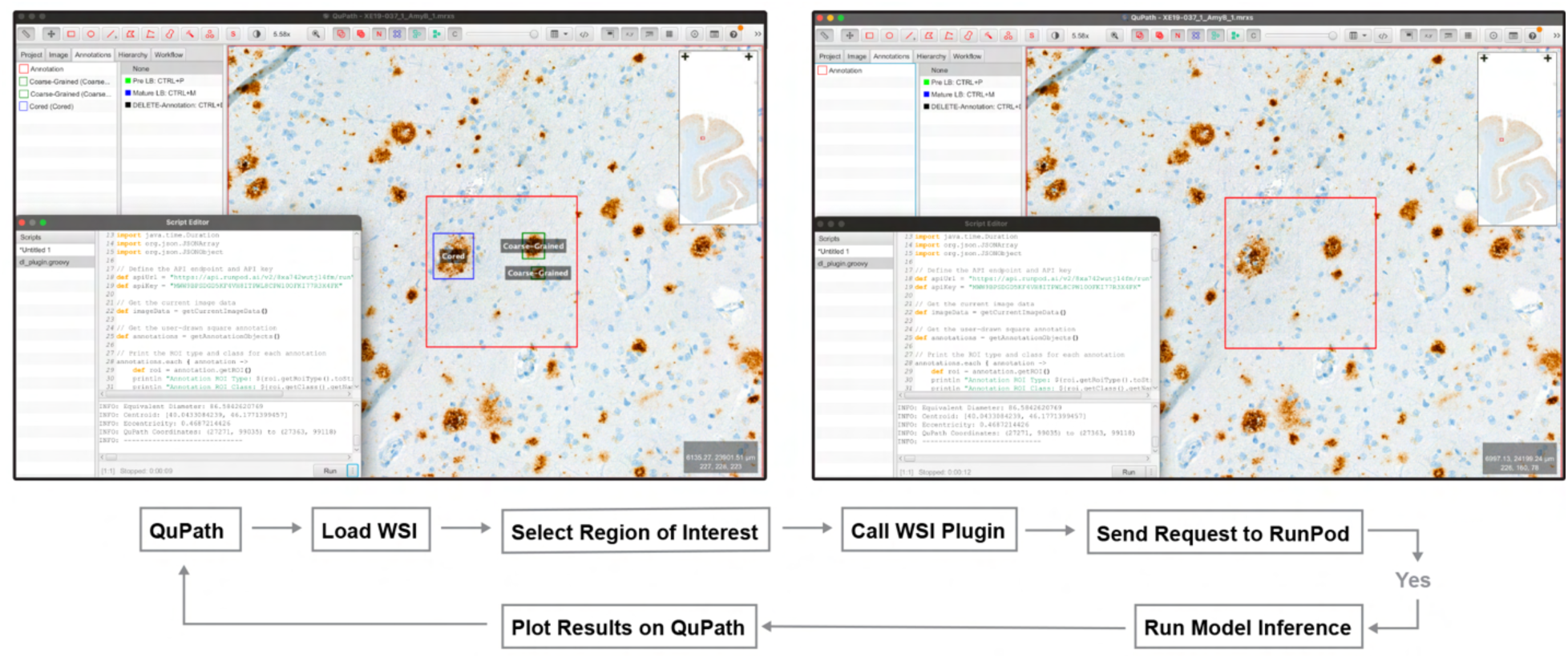
Deployment of AI model on QuPath. Demonstrates how user can use the trained model to generate predictions on real world data

## Supporting information

Supplemental Section

## Data Availability

All data produced in the present study are available upon reasonable request to the authors

## Acknowledgements

We wish to express our sincere gratitude to Dr. Brittany Dugger who generously made available digitized images of neuropathology of AD patients from the University of California, Davis Brain Bank for testing the generalizability of our algorithms. We would like to thank Dr. Laura Parkkinen for providing additional neuropathology input and expertise. We also wish to thank Françoise Chanut for editorial support and Kelley Nelson and Gayane Abramova for administrative support with manuscript preparation. This work was made possible primarily with support by the National Institutes on Aging (R01 AG067025) and the National Library of Medicine (R01 LM013617). Additional support was from the National Institutes of Neurological Disorders and Stroke (R01 NS124848) and the Koret Foundation Biomedical Research Program in Artificial Intelligence.

## Availability of Code

All the codes used in this study are publicly available in this git repository. https://github.com/finkbeiner-lab/amyb-plaque-detection.

