## Supplemental Section for "Generalizable Prediction of Alzheimer Disease Pathologies with a Scalable Annotation Tool and an High-Accuracy Model"

### Supplementary

| Feature | Dataset profile | Training Dataset (composed of train, validation, test WSI crops) |
|---|---|---|
| N (size of dataset) | 298 | 28 |
| Age | 81.5 +/- 10 years | 80.8+/- 10 years |
| Sex - female | 132 (44.3%) | 17 (60.7%) |
| DLB | 42 (14%) | 0 |
| AD | 146 (49%) | 28 (100%) |
| Dementia | 220 (73.8%) | 27 (96.4%) |
| No Dementia | 51(17.2%) | 0 |
| MCI | 27 (9%) | 1 (3.5%) |
| Control | 47 (15.7%) | 0 |
| PD | 10 (3.4%) | 0 |
| APOE - e4 | 69 (23.1%) | 7 (25%) |
| APOE - e2/e3 | 120 (40.3%) | 5 (17.8%) |

*DLB - dementia with Lewy bodies, PMI - Post Mortem Interval, AD - Alzheimer's Disease, MCI - Mild Cognitive Impairment, PD - Parkinson's Disease, APOE e4 - apolipoprotein E gene code ending with 4, APOE e2/e3 - apolipoprotein E gene code ending with 2 and 3*

**Supplementary Table 1:** Demographics of the Mt. Sinai dataset on which the study of plaque extraction and clinical correlation is performed

| **Attributes** | **Dataset 1- Mt. Sinai** | **Dataset 2 - UC Davis(External)** | **Dataset 3 UCD, UT, Univ of Pittsburgh (External)** |
|---|---|---|---|
| Total WSI's | 298 | 63 | 28 |
| Classes | Cored, Coarse-Grained, Diffuse, CAA | Cored, Diffuse, CAA | Cored, CAA |
| Center Crops | Objects are not center cropped | Objects are centered cropped | Objects are centered cropped |
| Tile Size | 1024 X 1024 | 256 X 256 | 256 X 256 |
| Magnification | x20 | x40 | x40 resized to x20 |
| Brain Regions | Middle Frontal Gyrus | Superior and Middle Temporal Gyrus | Temporal cortex |
| Antibodies | AB 4G8 | AB 4G8 | AB 4G8, NAB228, 6E10 |
| Staining | | chromagen 3,3′-diaminobenzidine and counterstained with hematoxylin | chromagen 3,3′-diaminobenzidine and counterstained with hematoxylin,<br><br>chromagen Nova Red from Vector (Sk-4800),<br><br>Leica Bond robotic immunostaining platform with proprietary detection reagents. Red chromogen which employs an alkaline phosphatase enzyme and a Fast Red chromogenic substrate |
| Thresholding for detecting candidate plaques for training | | | 4G8 HSV = (0, 40), (10, 255), (0, 220); NAB228 HSV = (0, 100), (1, 255), (0, 250); and 6E10 HSV = (0, 40), (10, 255), (0, 220) |

**Supplementary Table 2** Difference between various different external datasets The dataset varies in terms of total Whole Slide Images (WSI), classes, tile size, magnification, brain regions, antibodies and staining procedures.

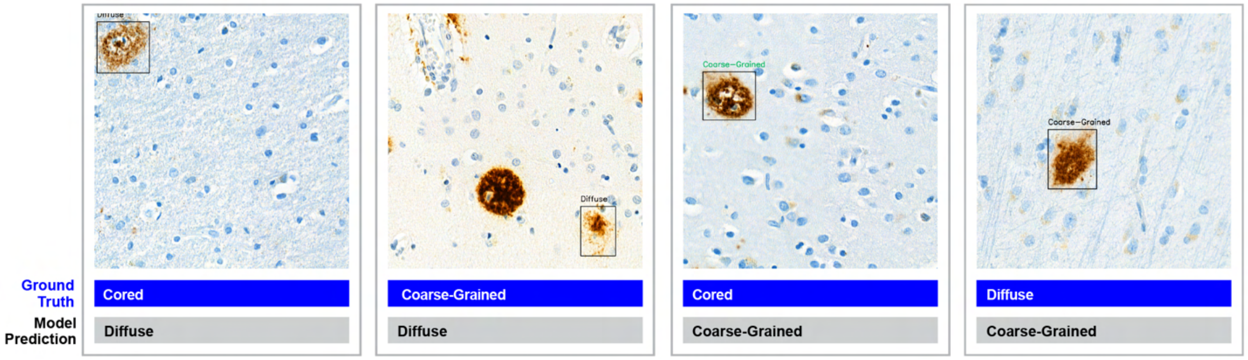


**Supplementary Figure 1**. Edge case examples where model gets confused with edge case examples

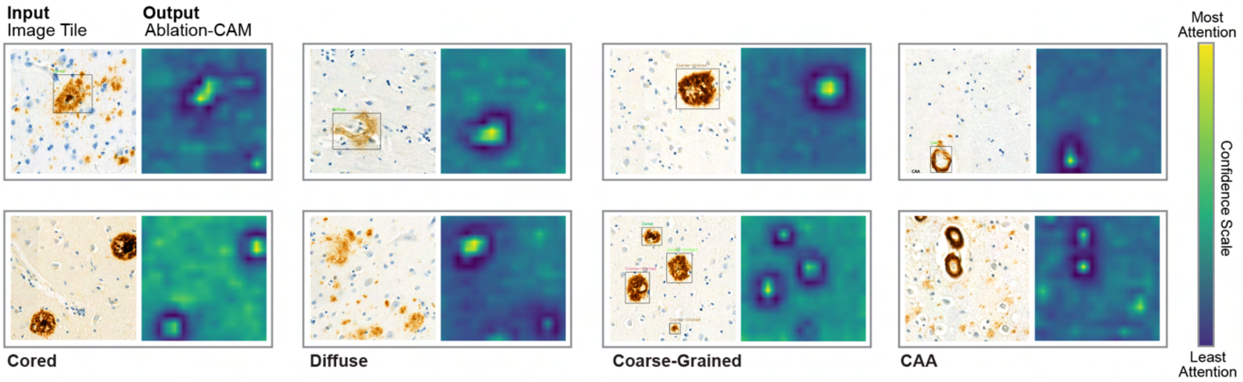


**Supplementary Figure 2**. Ablation CAM output. Analysis of the Mask R-CNN trained to identify and classify different classes of amyloid pathology with the Ablation-CAM tool to interpret AI reveals image features that the network learned to drive the classification.

| |
|---|
| Vertical Flip (p = 0.5) |
| HorizontalFlip (p = 0.5) |
| Blur(blur_limit = 3) |
| OpticalDistortion() |
| HueSaturationValue() |
| RandomRotate90() |

**Supplementary Table 3.** Data augmentation techniques from Albumentation library

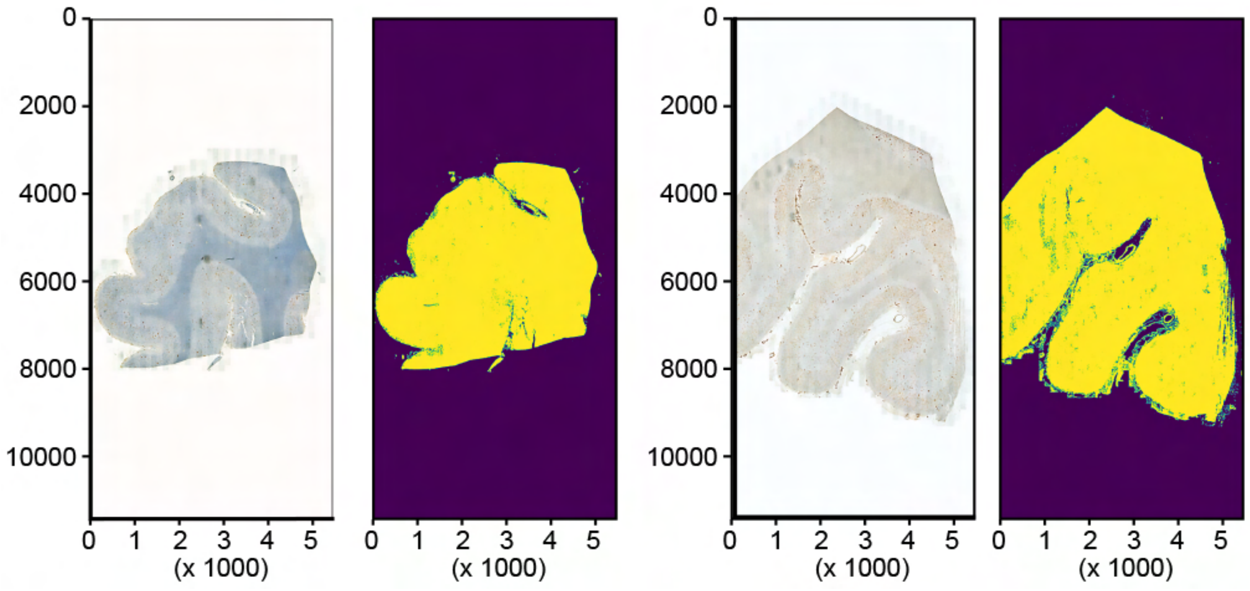


**Supplementary Figure 3**. WSI tissue segmentation from background using Otsu thresholding

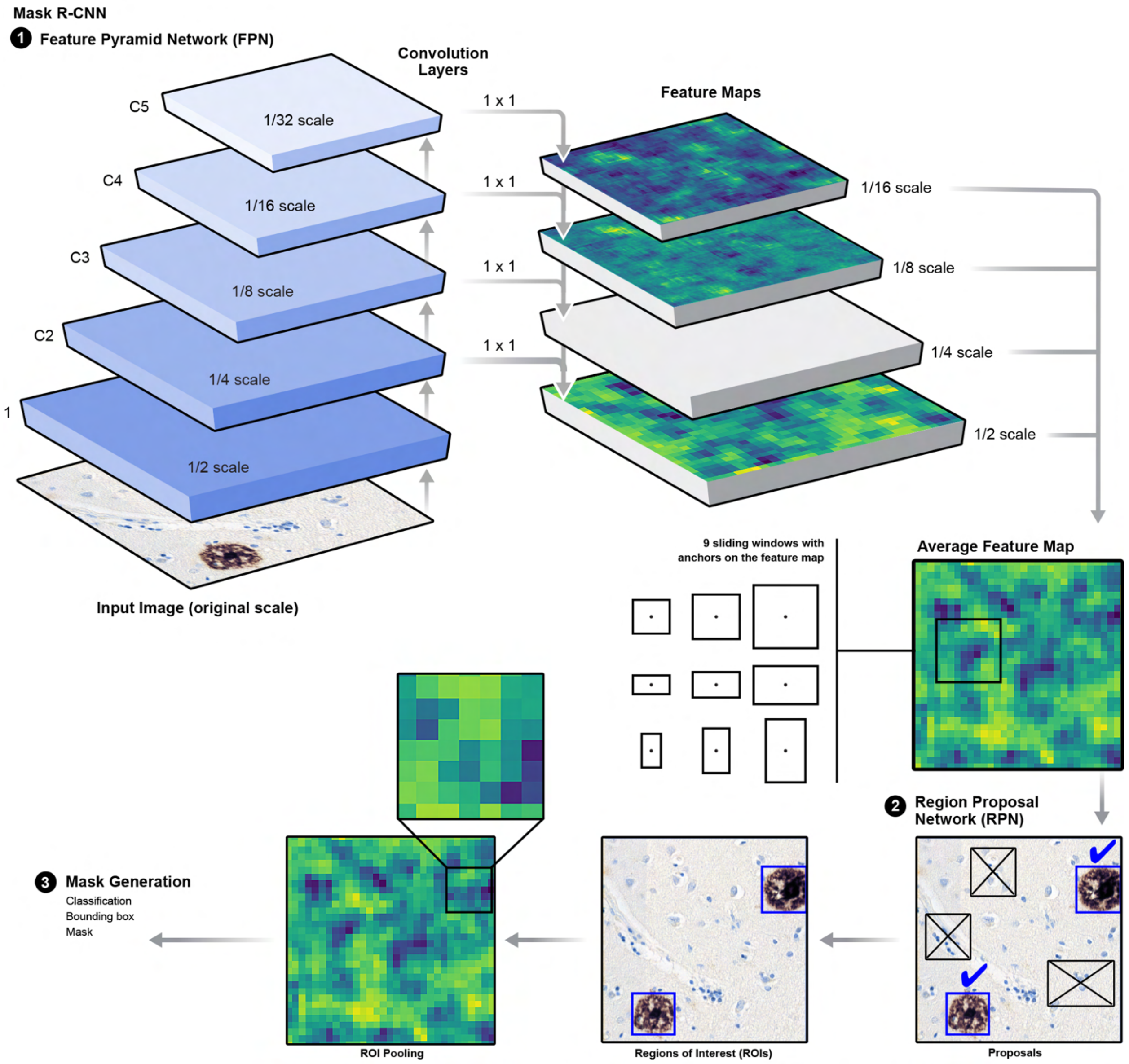


**Supplementary Figure 4**. Model architecture based on Mask R-CNN Feature Pyramid Network with ResNet50 as the backbone architecture. Feature maps are generated from images at different scales and are used for downstream prediction.

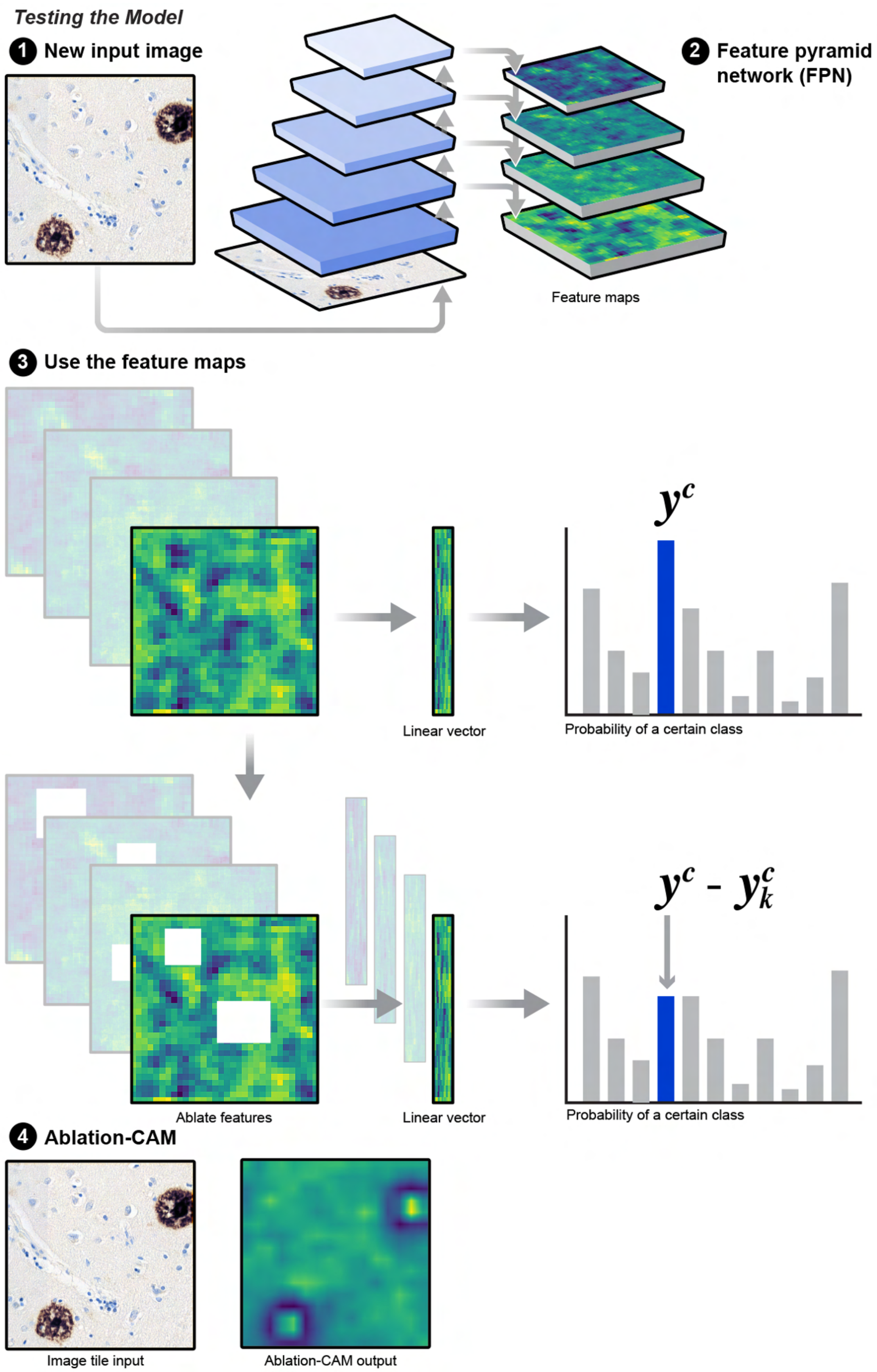

Testing the Model
1 New input image
2 Feature pyramid network (FPN)
Feature maps
3 Use the feature maps
$y^c$
Linear vector
Probability of a certain class
$y^c$ - $y^c_k$
Ablate features
Linear vector
Probability of a certain class
4 Ablation-CAM
Image tile input
Ablation-CAM output

**Supplementary Figure 5.** Model explainability using Ablation-CAM, illustrating each pixel's contribution to the prediction outcome."

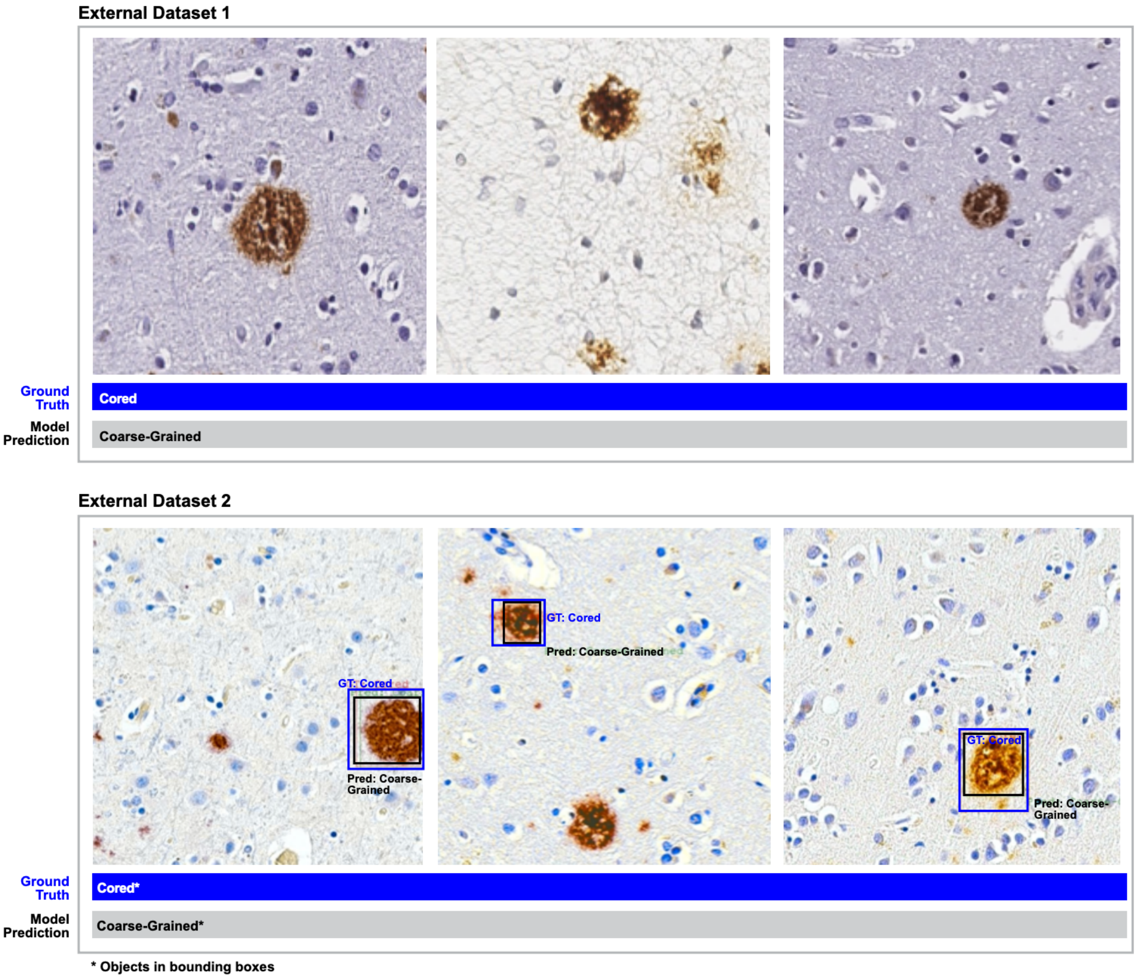


**Supplementary Figure 6:** The figure illustrates sample outputs for External Datasets 1 and 2, where the ground truth is labeled as "Cored" and the model prediction is "Coarse-Grained."
